# Impact of ambient temperature exposure on inflammation-related proteins: a repeated measurement study in the BAMSE cohort

**DOI:** 10.1101/2025.07.08.25331135

**Authors:** Zhebin Yu, Sophia Björkander, Annika Benndes, Federica Nobile, Jiawei Zhang, Simon Kebede Merid, Natalia Hernandez-Pacheco, Shizhen He, Maura M. Kere, Susanna Klevebro, Petter Ljungman, Massimo Stafoggia, Tom Bellander, Göran Pershagen, Anna Bergström, Inger Kull, Anne-Sophie Merritt, Niclas Roxhed, Olena Gruzieva, Jochen M. Schwenk, Erik Melén

## Abstract

**Background:** Short-term exposure to ambient temperature is linked to various health outcomes, raising public health concern in the context of climate change. We aimed to investigate longitudinal associations of temperature exposure with inflammation-related proteins among Swedish young adults.

**Methods:** We conducted three repeated measurements (2020-2022) by collecting self-sampled volumetric dry blood spots (DBS) from 807 participants from the Swedish BAMSE cohort (mean age 25.9 years). We estimated individual-address level daily temperature using a high-resolution spatiotemporal model. Inflammation-related proteins were measured using the Olink’s Explore Inflammation panel. Temperature-related proteins were identified using mixed-effect model adjusting for potential covariates, with potential effect modification by sex, smoking, asthma and air pollution explored. We further linked the temperature-related proteins to lung function, blood pressure and HbA1c. In addition, we built an inflammation-proteomic aging clock using a machine-learning approach and estimated the association between temperature exposure and proteomic age acceleration.

**Findings:** we found that 58 (16%) of the 365 studied inflammation-related proteins were significantly associated with short-term exposure to ambient temperatures (P<0.05 after correcting for multiple comparison). The impact of temperature exposure was modified by sex, smoking, asthma, and concurrent exposure to air pollution. A total of five, three and three temperature-related proteins were found to be associated with lung function, blood pressure, and HbA1c, respectively and validated in the UK Biobank. Peak temperature exposure (both cold and heat) was associated with significantly increased proteomic age acceleration.

**Interpretation:** Our findings suggest that ambient temperature exposure may cause adverse health effects through perturbating inflammation-related proteins.

**Synopsis:** This study reports significant effects of ambient temperature exposure on inflammation-related proteins, highlighting potential health impacts from ambient temperature exposure.

**Graphical abstract:** 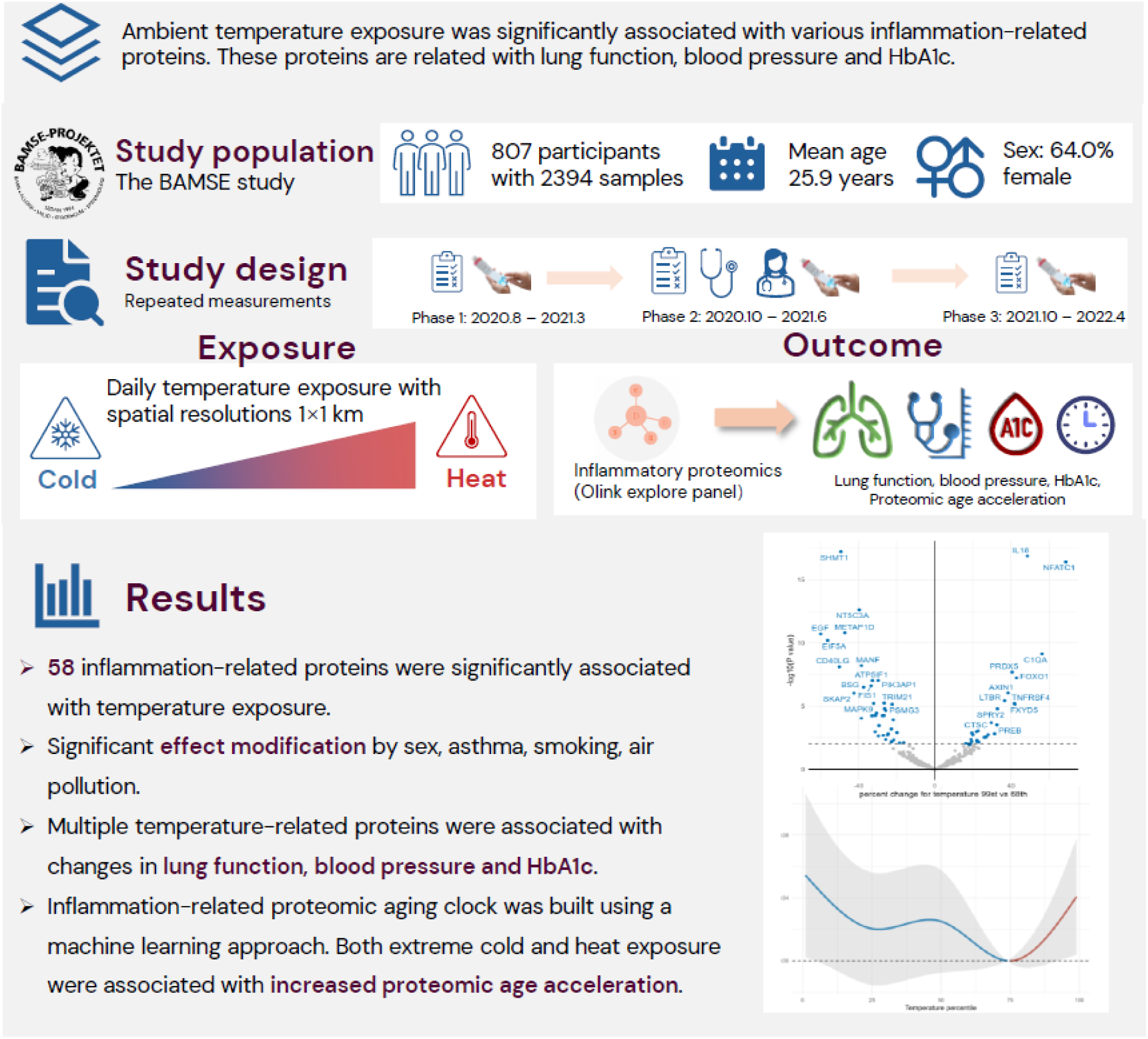

## Introduction

Numerous studies have shown that non-optimum temperature exposure (both extreme heat and cold) is associated with a significant disease burden of morbidity and mortality^1–3^. The frequency of extreme heat and cold events is likely to increase in the context of climate change^4,5^. However, the potential mediators and mechanisms underlying the health impacts of non-optimum temperature exposures still need to be disentangled. Exploring mechanisms can help us understand how temperature change can trigger new diseases, exacerbate existing diseases, and influence diagnostic and therapeutic approaches. Such knowledge is critically important to develop appropriate resilience and recovery strategies and health interventions towards climate change^6^.

Inflammation is one potential temperature-sensitive disease mechanism due to its crucial role in the body’s response to tissue damage and injury. Previous studies have explored the association between temperature exposure and inflammation biomarkers^7–12^. For example, Cheng et al.^7^ used 80,574 electronic health examination records to show that short-term exposure to heat waves was associated with increased systemic inflammation markers, including white blood cell- and neutrophil counts. Ni et al.^8^ reported cross-sectional associations between cold exposure and subclinical inflammation markers in the KORA F4 study. However, these studies were limited by a relatively low number of targeted biomarkers, cross-sectional design (with difficulty controlling confounding), or a small sample size. Given the complexity of inflammatory processes, there is a need for more studies to capture a broader range of inflammatory markers to gain a more comprehensive picture of the impact of non-optimal temperature on health.

In the present study, we aimed to investigate the associations between temperature exposure and 365 inflammation-related proteins in longitudinally self-sampled blood from a Swedish population-based cohort. Additionally, we explored factors that might modify the association of peak temperature with proteins and evaluated the association of the significant proteins with cardiometabolic and pulmonary phenotypes. Finally, we built an inflammation-related proteomic aging clock and linked that to the temperature exposure to gain more insights into overall health impacts.

## Methods

### Study population

Data from the BAMSE (Children, Allergy, Milieu, Stockholm, Epidemiology) study, which an ongoing population-based birth cohort originally comprising 4089 children born between 1994 and 1996, was used for this study. Participants have been followed up with repeated questionnaires and clinical examinations up to adulthood (a 24-year follow-up was finished in 2019^13,14^). During the pandemic, specific COVID-19 follow-ups^15,16^ with web-based questionnaires and a clinical examination were further conducted (Phase 1 from August 2020 to March 2021, Phase 2 from October 2020 to June 2021, Phase 3 from October 2021 to February 2022, **Supplemental Figure 1**). Quantitative dry blood spot (qDBS, Capitainer AB, Sweden) samples were collected from finger pricking^17,18^ (self-sampled at home in Phase 1 and Phase 3, self-sampled at the clinic in Phase 2, **Supplemental Figure 1**). The current analysis was based on 808 participants who contributed blood samples at Phase 1-3 of the COVID-19 follow-up. After exclusion of 30 samples due to sampling or assay errors, a total of 2394 samples belonging to 807 participants (Phase 1 n=798, Phase 2 n=793, Phase 3 n=803) were included and 783 subjects had three consecutive measurements (**Supplemental Figure 1**). This study was approved by the Swedish Ethical Review Authority (approval 2020-02922), and all participants gave written informed consent.

### Measurement of inflammation-related proteins

The qDBS samples were eluted in 100 µl of elution buffer (1x Phosphate-buffered saline (PBS, Medicago) with 0.05% Tween 20 and protease inhibitor cocktail (#04693116001, Roche)) for 60 minutes at room temperature under gentle rotational shaking (170 r.p.m). Inflammation-related proteins in the DBS eluates were then measured by proximity extension assays (PEA) at the SciLifeLab Affinity Proteomics Unit in Uppsala using Olink’s Explore 384 Inflammation panel^19^. The quantification of relative protein levels was reported as normalized protein expression (NPX) arbitrary units on a log2 scale. Protein-specific probabilistic quotient normalization (ProtPQN)^20,21^ was further applied to the protein levels to account for non-biological differences in DBS obtained from self-sampling.

### Exposure assessment

Daily average ambient temperature exposure with spatial resolution 1×1km was derived from a machine learning model^22^ (Land Use Random Forest, LURF) covering Sweden. In brief, satellite land surface temperature (LST) was combined with modeled air temperature from atmospheric models to generate complete LST data. The imputed LST data and monitored air temperature data were calibrated using spatial parameters (land use variables, climatic zones, population density, elevation, normalized difference vegetation index (NDVI), and meteorological variables). Ten-fold cross-validation was performed with R^2^=0.94, root mean squared error=1.6 ℃ on average across the years. Information on participants’ residential history up to 2023 was collected based on questionnaire reports and updated using Swedish Tax Agency records. We further assigned daily air temperature exposure to each participant’s corresponding residential address at the biosampling dates. Daily exposure to fine particulate matter (PM_2.5_), nitrogen dioxide (NO_2_), ozone (O_3_), and relative humidity were derived from an urban background monitoring station in central Stockholm^23^ .

### Cardio-metabolic and pulmonary phenotypes

During the clinical examinations of Phase 2 COVID-19 follow-up, lung function was measured using a Vyaire Vyntus spirometer according to the European Respiratory/American Thoracic Society guidelines^24^ to determine forced expiratory volume in 1s (FEV1), forced vital capacity (FVC), and the ratio FEV1/FVC. Blood pressure was measured using the Omron HBP-1300 Automatic monitor following the standard operating procedure, including quality control measures. Each participant was assessed three times with one minute rest between the measurements, and the average blood pressure values are used in the current study. The levels of hemoglobin A1c (HbA1c) were quantified in non-fasting plasma samples using capillary electrophoresis at the accredited Karolinska University Hospital Laboratory (Stockholm, Sweden), and reported in mmol/L according to the international HbA1c calibration (IFCC).

### Covariates

Covariates were derived from the questionnaires at the 24-year follow-up and COVID-19 follow-ups. Body mass index (BMI) was calculated using the height and weight measured at Phase 2. Current smoking was defined as positive answer to the question “Do you smoke?” at any phase of the COVID-19 follow-up. Asthma was defined based on at least two of the three following criteria in the last 12 months before the Phase 1: (1) symptom of wheezing; (2) ever having a doctor’s diagnosis of asthma; (3) use of asthma medication^25^. History of SARS-CoV-2 infection was determined for each phase using a cumulative approach, based on the following criteria: (1) positive results for SARS-CoV-2 polymerase chain reaction (PCR) testing up to the date of biosampling, obtained through linkage to the Swedish registry of infectious diseases (SmiNet); (2) positive serology based on the presence of SARS-CoV-2 antibodies in either serum (Karolinska University Laboratory) or in plasma as described in Björkander et al^16^ measured during Phase 2 ; and (3) self-reported positive results from PCR, antibody, or antigen tests, as reported in the questionnaires for Phase 1-3^15^. COVID-19 vaccination status up to the sampling dates of each participant was determined based on linkage to the national vaccination register^26^.

### Inflammation-related proteomic aging clock

We applied the light Gradient Boosting Machine (lightGBM) to use all included inflammation-related proteins to predict the chronological age. The chronological age was calculated based on the exact sampling date at each phase and birth date. To account for the repeated-measure structure of the data, we first ran linear mixed models using each protein level as outcome and individual as random intercept. The residuals of the linear mixed models were further used as predictors in the lightGBM model. We split the samples into training and testing datasets, consisting of 70% and 30% of the study population, respectively. In the training dataset, the model was tuned based on five-fold cross-validation to maximize the R-square, and feature selection was conducted using Boruta with 500 iterations, which selected 47 proteins relevant to the chronological age. After training the model, the final lightGBM model was fitted using the final hyperparameters, and protein-predicted age values were calculated for the whole dataset. Finally, proteomic age acceleration was calculated based on protein-predicted age minus chronological age.

### Statistical analysis

We employed a linear mixed model with an individual-specific random intercept to account for the repeated-measurement design. To explore both the lagged exposure time window and non-linear associations, we applied the distributed lag nonlinear model (DLNM) for the ambient temperature at 0-4 days before the blood draw. The DLNMs used ‘cross-basis’ function combining the lag-response and exposure-response, which is particularly useful to estimate the lagged and non-linear effect of environmental exposure. We applied the nature splines with three knots placed at 25th, 50th and 75th percentiles for the exposure-response curve as well as three internal knots equally distributed for the lag-response curve. Due to the distribution of protein levels significantly different from normality, the results of the ProtPQN analysis were normalized using the rank-based inverse normal transformation method. The output of this step was used as the dependent variable of the model. Covariates included in the mixed model were determined *a priori* and included: age, sex, long-time trend (nature spline term with six degrees of freedom for the whole study period), follow-up phases, day of the week, season (warm: April to September; cold: October to March), history of SARS-CoV-2 infection, and vaccination at the time of follow-ups. We first tested whether the ambient temperature exposure significantly contributed to the variations of the protein levels by comparing the models (protein levels as outcomes and all covariates as independent variables) with and without temperature using the likelihood test. P-values were corrected for multiple comparisons using the Benjamini-Hochberg procedure. Secondly, the relative “cold” and “heat” effect estimates on specific proteins were further quantified by comparing the 1^st^ percentile (-9.6 ℃) and 99^th^ percentile (21.0 ℃) exposures with the reference temperature. In the current study, we selected the most frequent temperature during the study period as the reference temperature^27^ (68^th^ percentile of the temperature distribution equals 15 ℃), and in the sensitivity analysis we also tested the other reference temperature including median and 25^th^ 75^th^ percentiles.

Subgroup analyses were conducted to test whether the effect size of the association of temperature with blood proteins differed by sex, asthma status, active smoking, and short-term air pollution exposure levels (medium air pollution level as the cut-off), with difference assessed using the formula: 𝛽_1_ − 𝛽_2_ ± 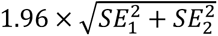, where β_1_ and β_2_ were the coefficients for two subgroups and SE_1_ and SE_2_ were the corresponding standard errors We carried out the following sensitivity analyses to test the robustness of our findings: 1) Using the cumulative mean temperature for lag0-4 days as exposure without DLNM; 2) Using different lag days of temperature (lag0-1, lag0-2, lag0-3, lag0-5, lag0-6, lag0-7, lag0-14); 3) Changing the referent temperature to 50^th^ percentile or 75^th^ /25^th^ percentile (for heat or cold effect, respectively); 4) Additionally adjusting for PM_2.5_, NO_2_, O_3_, and relative humidity on lag 0-1 in separate models; 3) Restricting the analysis to Phase 1 and 2, or Phase 2 and 3.

The significant temperature-related proteins were further tested in association with spirometry measurements (pre-bronchodilator FEV_1_, FVC, FEV_1_/FVC), systolic/diastolic blood pressure (SBP/DBP), and HbA1c using linear regression models with adjustment for age, sex, season, and height (for lung function) or BMI (for blood pressure and HbA1c). The significant associations of specific proteins with the same phenotypes were attempted for replication in the UK Biobank Pharma Proteomics Project (UKB-PPP)^28^. Because association results with spirometry measurements were not available among the publicly available UKB-PPP results^28^, we tested the associations between proteins with respiratory diseases (ICD-10 code J00-J99) instead given the well-documented relationships between lung function and chronic airway diseases. Evidence of replication was considered for significant proteins after adjusting for multiple comparisons (Benjamini-Hochberg procedure) and with the same direction of the association effect with the phenotype evaluated in the UKB-PPP (British or Irish ancestry, N=46,218).

The association between temperature exposure and inflammation-related protein-based age acceleration was estimated using the linear mixed model with the same adjustment as the temperature-protein association analysis. The reference temperature was first selected based on the corresponding temperature of the lowest proteomic aging acceleration and then the exposure-response curve was plotted. Protein-Protein Interaction (PPI) networks were investigated using the STRING (v.12) online tool. All analyses were conducted using the FRGEpistasis, lme4, splines, dlnm, lightGBM and Boruta packages implemented in the R statistical software, version 4.3.2 (The R Foundation for Statistical Computing, Vienna, Austria).

## Results

A total of 2,394 DBS samples (Phase 1, n=798; Phase 2, n=793; Phase 3, n=803) were included in the analysis, with 64.5% female donors and a mean age of 25.9 ± 0.95 years. The basic clinical and demographic characteristics of the participants across the three phases are presented in **Table 1**. Compared with all the participants of the most recent BAMSE cohort 24-year follow-up, the included subjects were more often female, had higher education, and fewer were smokers but had similar distributions in age, occupation, BMI, and asthma status (**Supplemental Table 1**).

**Table 1.** Clinical and demographic characteristics of the study population across three data collection phases.

|  | Phase 1 (N=798) | Phase 2 (N=793) | Phase 3 (N=803) |
| --- | --- | --- | --- |
| Sampling |  |  |  |
| Collection start | August 2020 | October 2020 | October 2021 |
| Collection end | March 2021 | June 2021 | April 2022 |
| Female | 511 (64.0%) | 509 (64.2%) | 512 (63.8%) |
| Age (years) | 25.3 (0.78) | 25.8 (0.82) | 26.6 (0.78) |
| Smoking | 131 (16.4%) | 131 (16.5%) | 129 (16.1%) |
| Asthma | 128 (16.0%) | 126 (15.9%) | 128 (15.9%) |
| History of SARS-CoV-2 infection | 49 (5.9%) | 271 (30.5%) | 355 (32.8%) |
| COVID-19 vaccination |  |  |  |
| 1st dose | 0 (0%) | 24 (3.0%) | 748 (93.2%) |
| 2nd dose | 0 (0%) | 12 (1.5%) | 730 (90.9%) |
| 3rd dose | 0 (0%) | 0 (0%) | 28 (3.5%) |
| Environmental Exposure |  |  |  |
| Temperature, °C | 13.0 (3.0) | 5.6 (6.8) | 3.2 (5.8) |
| PM2.5, µg/m<sup>3</sup> | 3.2 (2.2) | 5.0 (3.3) | 6.6 (6.0) |
| NO<sub>2</sub>, µg/m<sup>3</sup> | 7.2 (1.5) | 11.3 (5.7) | 12.6 (7.7) |
| Ozone, µg/m<sup>3</sup> | 57.0 (10.9) | 51.8 (18.2) | 41.2 (11.8) |
| Relative humidity, % | 62.5 (7.8) | 75.9 (15.3) | 84.9 (8.7) |
| Cardiometabolic and pulmonary phenotypes |  |  |  |
| FEV1, L | - | 3.9±0.8 | - |
| FVC, L | - | 4.8±1.0 | - |
| FEV1/FVC, % | - | 82.8±6.0 | - |
| SBP, mmHg | - | 111.2±10.2 | - |
| DBP, mmHg | - | 94.6±8.5 | - |
| HbA1c, mmol/mol | - | 31.5±2.5 | - |

Data are presented as frequency (percentage) for categorical variables and mean (standard deviation, SD) for continuous variables.501 Abbreviations: PM2.5, particulate matter with diameter $\leq 2.5$ µm; NO<sub>2</sub>, nitrogen dioxide; NO<sub>x</sub>, nitrogen oxides; FEV1, Forced Expiratory Volume in one second; FVC, Forced Vital Capacity; SBP, Systolic Blood pressure; DBP, Diastolic Blood Pressure; HbA1c, Hemoglobin A1c;

The mean temperatures at the self-sampling dates were 13.0±3.0 ℃, 5.6±6.8, ℃ and 3.2±5.8 ℃ at each of the three phases, respectively. We observed moderately negative correlations between NO2 (r= -0.62), humidity (r=-0.53), and ambient temperature, whereas a moderately positive correlation between ozone and temperature (r=0.44), and a low correlation with PM2.5 were found (r = 0.13, **Supplemental Figure 2**). We identified 58 out of 365 inflammation-related proteins significantly associated with ambient temperature exposure after adjusting for potential covariates and correcting for multiple comparisons (**Table 2, Supplemental Table 2**). The exposure-response curves between temperature and proteins are presented in **Supplemental Figure 3**. The association effect estimates of exposure to cold (1^st^ percentile) and “heat” (99^th^ percentile) compared with the reference temperature (68^th^ percentile) for each protein are presented in **Figures 1 A and B**. In brief, cold exposure was significantly associated (FDR < 0.05) with 48 proteins, of which 34 showed a positive direction of the effect size, and association with lower levels was observed for 14 proteins. In comparison, “heat” exposure was associated with five proteins after FDR adjustment. Since exposure-response relationships for the five proteins significantly associated with heat exposure were monotonic (**Supplemental Figure 3**), only the estimates for low temperature exposure were presented for the subsequent subgroup and sensitivity analysis. The association estimates for cold exposure remained similar across sensitivity analyses, including different exposure lags, additionally adjusting for air pollution exposure, excluding participants taking asthma medications or restricted to certain phases of follow-ups (**Figure 1 C and Supplemental figure 4**).

**Figure 1.**
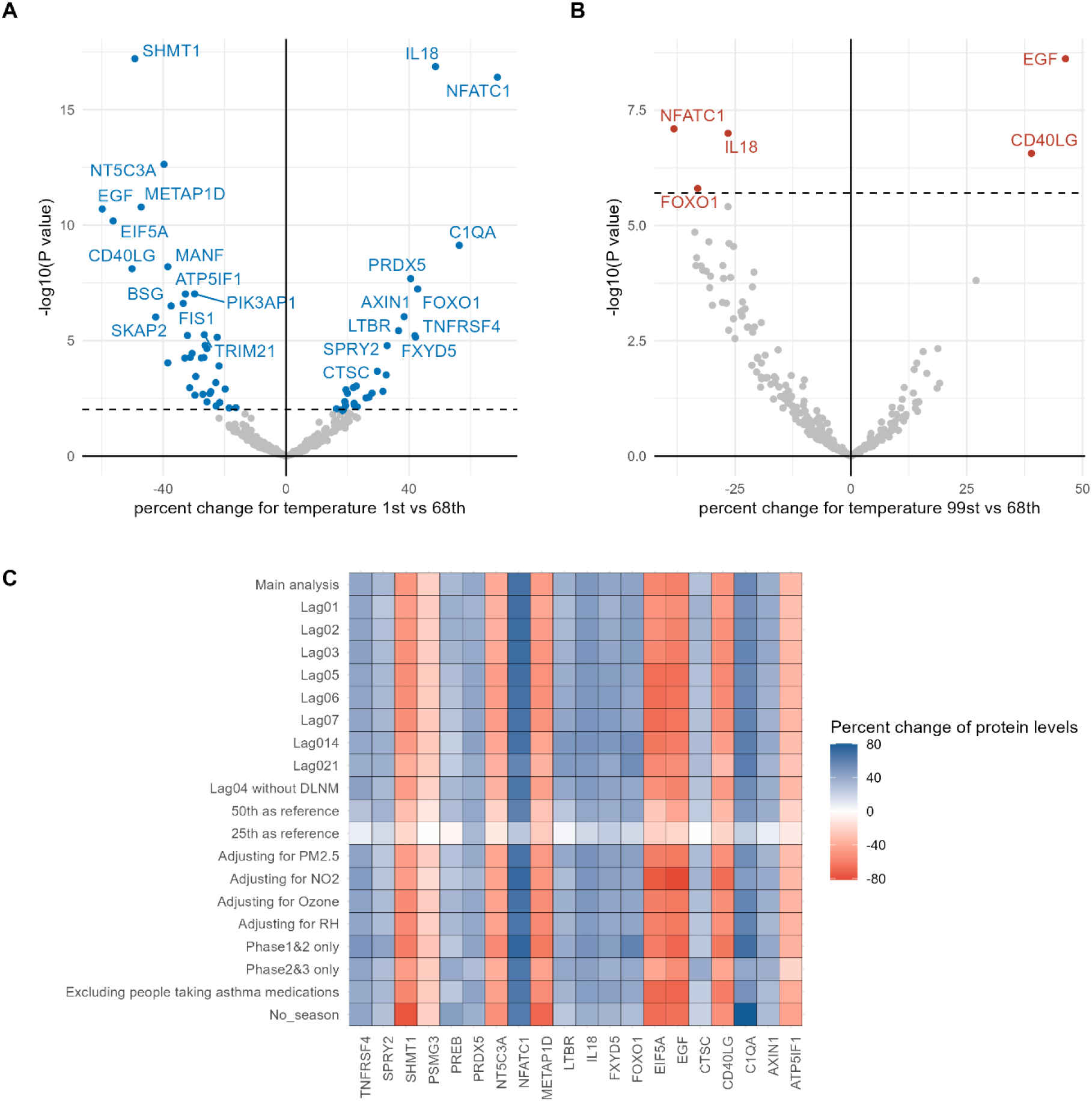
Associations between daily ambient temperature and inflammation-related proteins. Volcano plots show the associations between (A) extreme low (1^st^ percentile) and (B) extreme high (99th percentile) temperature and inflammation-related protein levels compared with the reference temperature (68^th^ percentile). Results are shown in terms of the logarithmic transformation of the p-value (-log_10_ p-value) on the *y*-axis and coefficient estimates with temperature exposure on the *x*-axis. Proteins significantly associated with cold or heat after correcting for the False Discovery Rate (FDR) are illustrated by blue or red dots, respectively. Dashed horizontal lines indicate the FDR significance level. (C) Association effect estimates with cold temperature exposure for the top 20 most significant proteins in the sensitivity analysis. All the results were adjusted for age, sex, long time trend (nature spline term with six degrees of freedom), phases, day of the week, season (warm: April to September; cold: October to March), history of SARS-CoV-2 infection and vaccination at the time of biosampling.

**Table 2.**
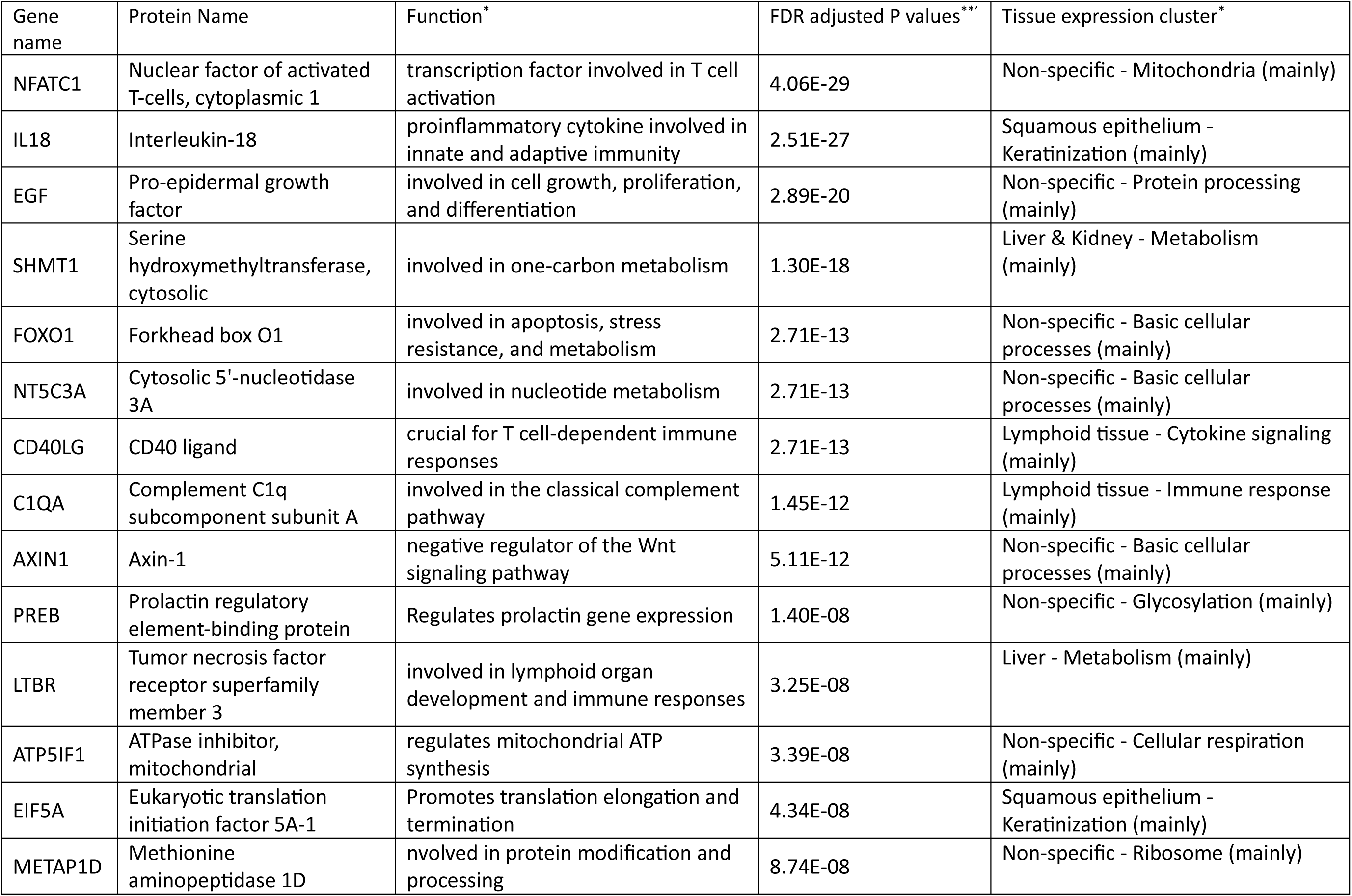
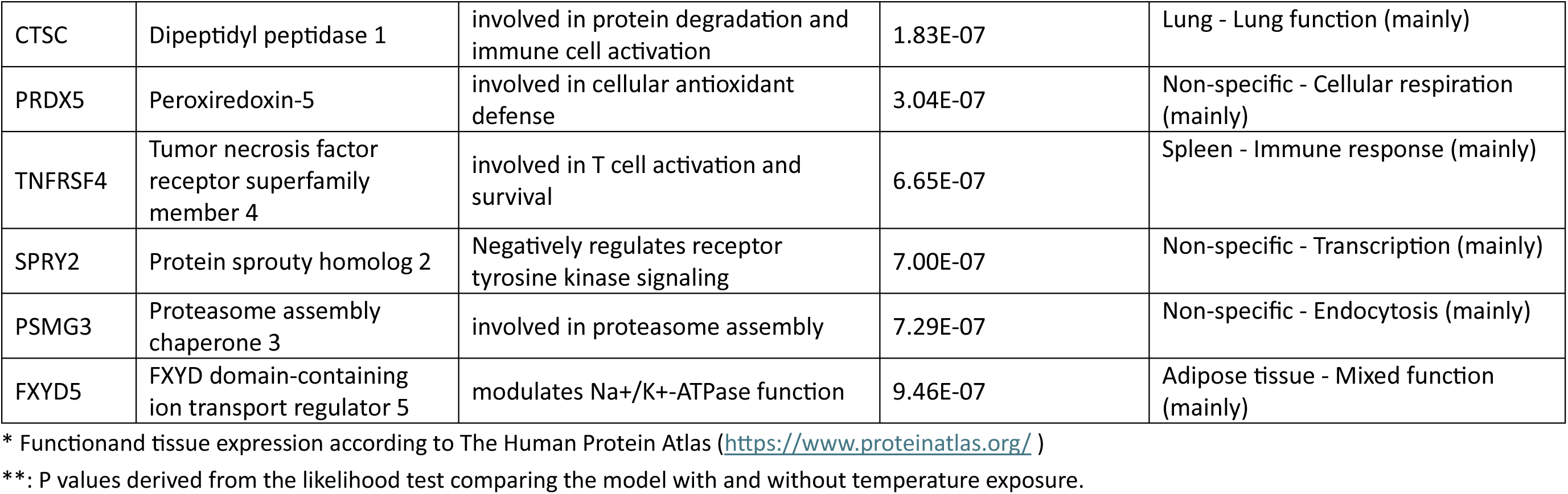
Top 20 proteins significantly associated with ambient temperature exposure.

After stratifying the analysis by sex, we observed that the effect of cold exposure was significantly stronger in males for nine proteins (ADGRE2, CTSC, FXYD5, LAP3, NBN, PREB, SPINT2, SPRY2, TNFRSF13B) and in females for three proteins (SKAP2, EIF5A, CXCL1) (**Figure 2**). Additionally, we found that the association effect estimates of cold were significantly higher among participants with asthma compared with non-asthmatic participants for four proteins (AGRE2, CTSC, EIF5A, NBN). We also observed positive associations between cold exposure and CSF1 and DNER only among non-smokers. For air pollution, we observed that the cold effect estimates were significantly different for six proteins when exposed to different air pollution levels (BCR, ANGPTL2 for PM2.5; METAP1D, LAMP3, CSF1, ATP5IF1 for NO2).

**Figure 2.**
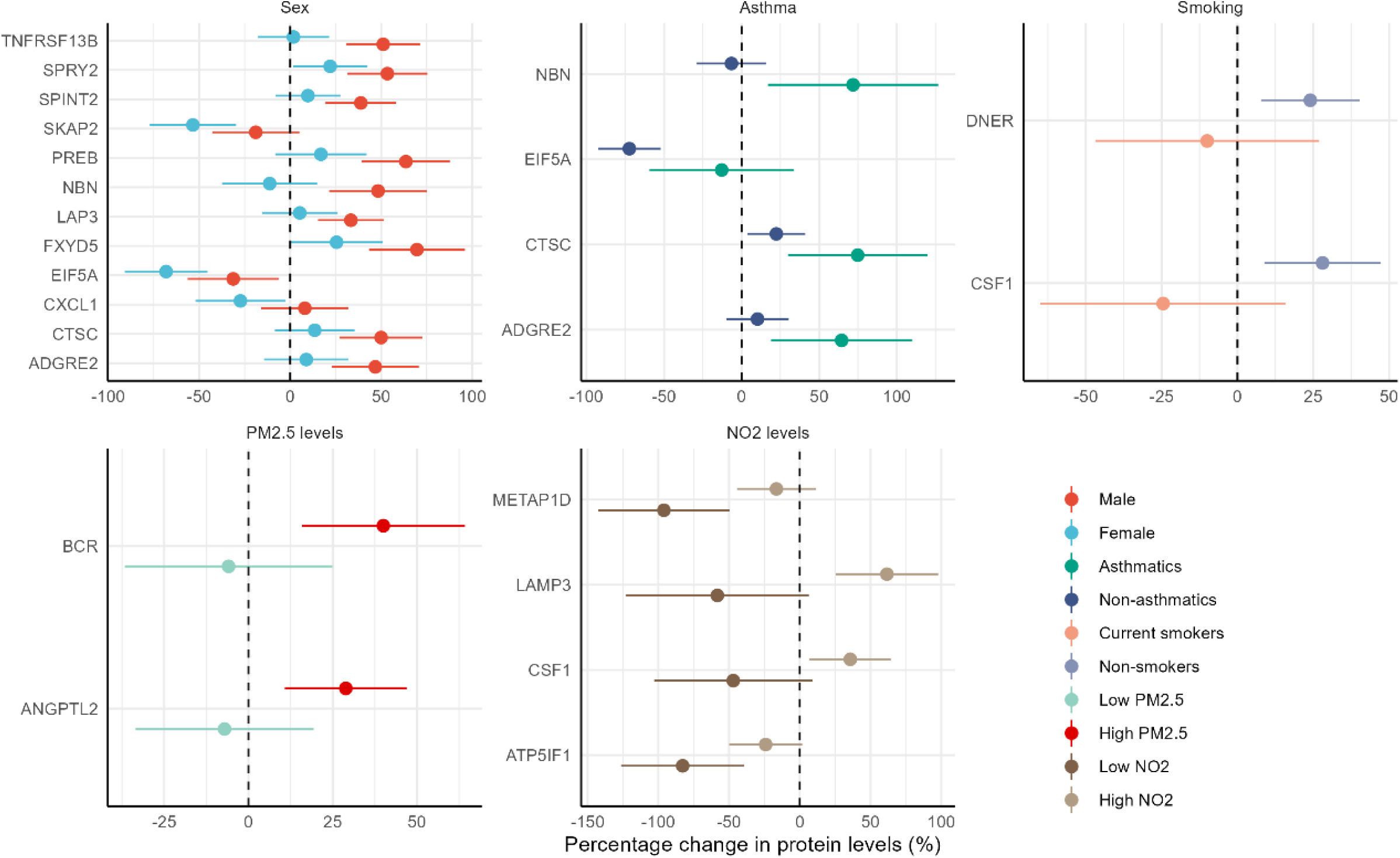
Association results between daily ambient temperature and inflammation-related protein levels stratified by sex, asthma status, current smoking, and air pollution levels. All the results were adjusted for age, sex, long time trend (nature spline term with six degrees of freedom), phases, day of the week, season (warm: April to September; cold: October to March), history of SARS-CoV-2 infection and vaccination at the time of biosampling.

Among the temperature-related proteins, we found 12, 11, and eight proteins associated with FEV1, FVC, and FEV1/FVC measured in the Phase 2 clinical examination. Additionally, several proteins were associated with SBP, DBP, and HbA1c (**Figure 3**). The overlapping significant proteins among different phenotypes are presented in **Supplemental Figure 5**. Among these, TNFRSF4 was found to be associated with both lung function, blood pressure, and HbA1c levels. Moreover, five proteins were significantly associated with chronic respiratory diseases in UKB-PPP, as a proxy of lung function, namely DNER, TGFB1, TNFRSF13B, TNFRSF4, SCGB1A1. The association with blood pressure (NFATC1, TNFRSF4, SCGB1A1) and HbA1c (C1QA, MEPE, SCGB1A1) was also replicated (**Supplemental Table 3**). The PPI network analysis showed complex interactions (enrichment *P*-value=3.3e-12) with the top significant proteins as the key nodes such as NFATC1, IL18, and CD40 (**Supplemental Figure 6**).

**Figure 3.**
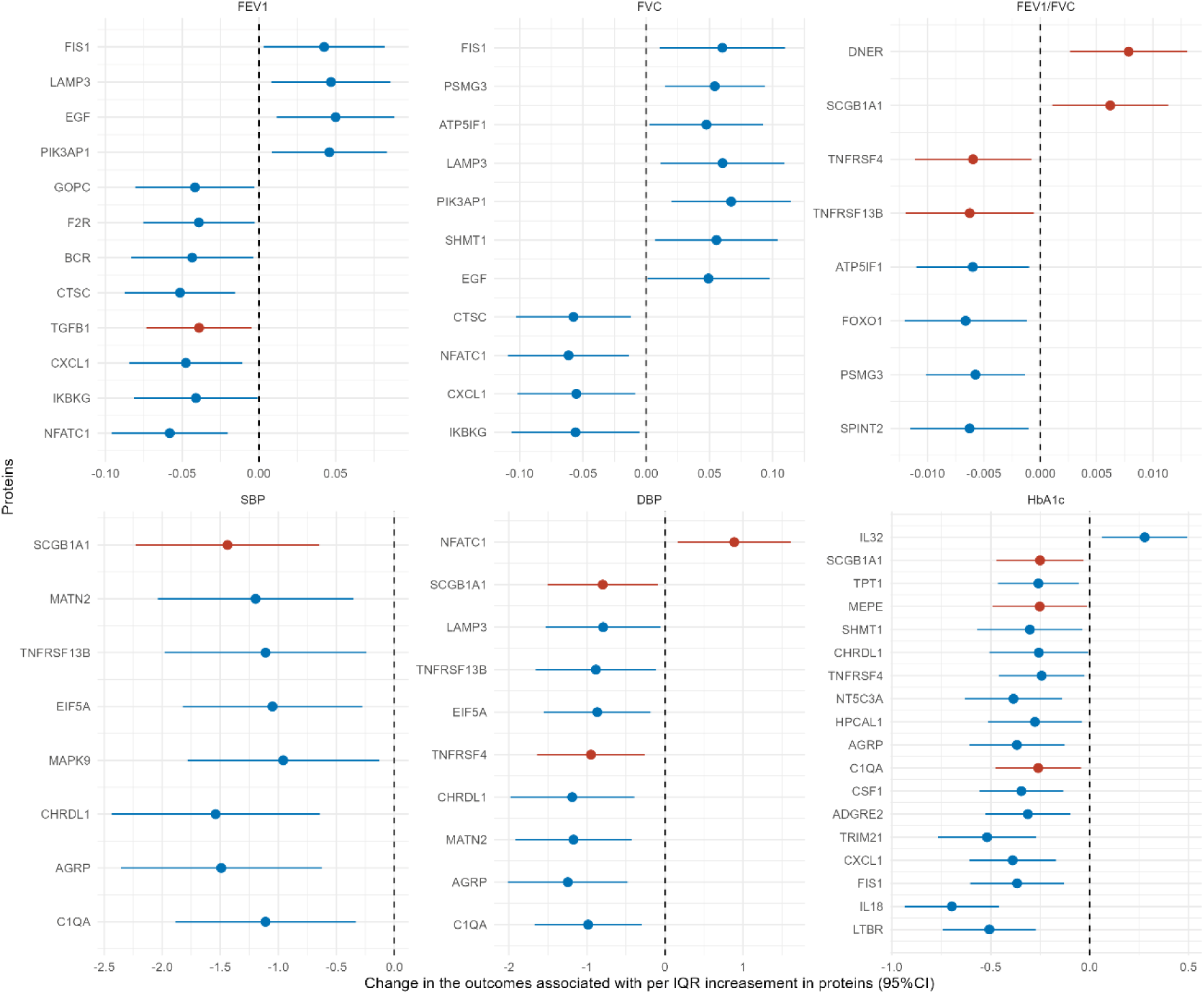
Association between significant inflammation-related proteins and cardiometabolic and pulmonary phenotypes. The y-axis shows the changes in the phenotypes associated with per interquartile range increase in the protein levels with the bar indicating 95% confidence intervals. Only the inflammation-related proteins that were significantly associated with temperature exposure were presented in the figure. Proteins with red dots indicating the association for these proteins also observed in the UK biobank Pharma Proteomic Project with consistent direction of the association effect. Results were adjusted for age, sex, height (for FEV1, FVC, FEV1/FVC as outcome), or body mass index (SBP, DBP, HbA1c as outcome). FEV1, Forced Expiratory Volume in one second; FVC, Forced Vital Capacity; SBP, Systolic Blood pressure; DBP, Diastolic Blood Pressure; HbA1c, Hemoglobin A1c

The inflammation-related proteomic age, predicted using 47 proteins, showed a high correlation with chronological age (Pearson correlation 0.93 and R-square 0.85, **Figure 4A**), with RABGAP1L identified as the most important protein in the lightGBM model (**Figure 4B**). The distribution of proteomic age acceleration was similar across three phases (**Figure 4C**). The exposure-response curve between temperature exposure and proteomic age acceleration showed that both extreme heat and extreme cold were associated with a significant proteomic age acceleration compared with the reference temperature (0.04 (95%CI:0.02-0.09) and 0.05 (95%CI:0.03-0.10), respectively, **Figure 4D**).

**Figure 4.**
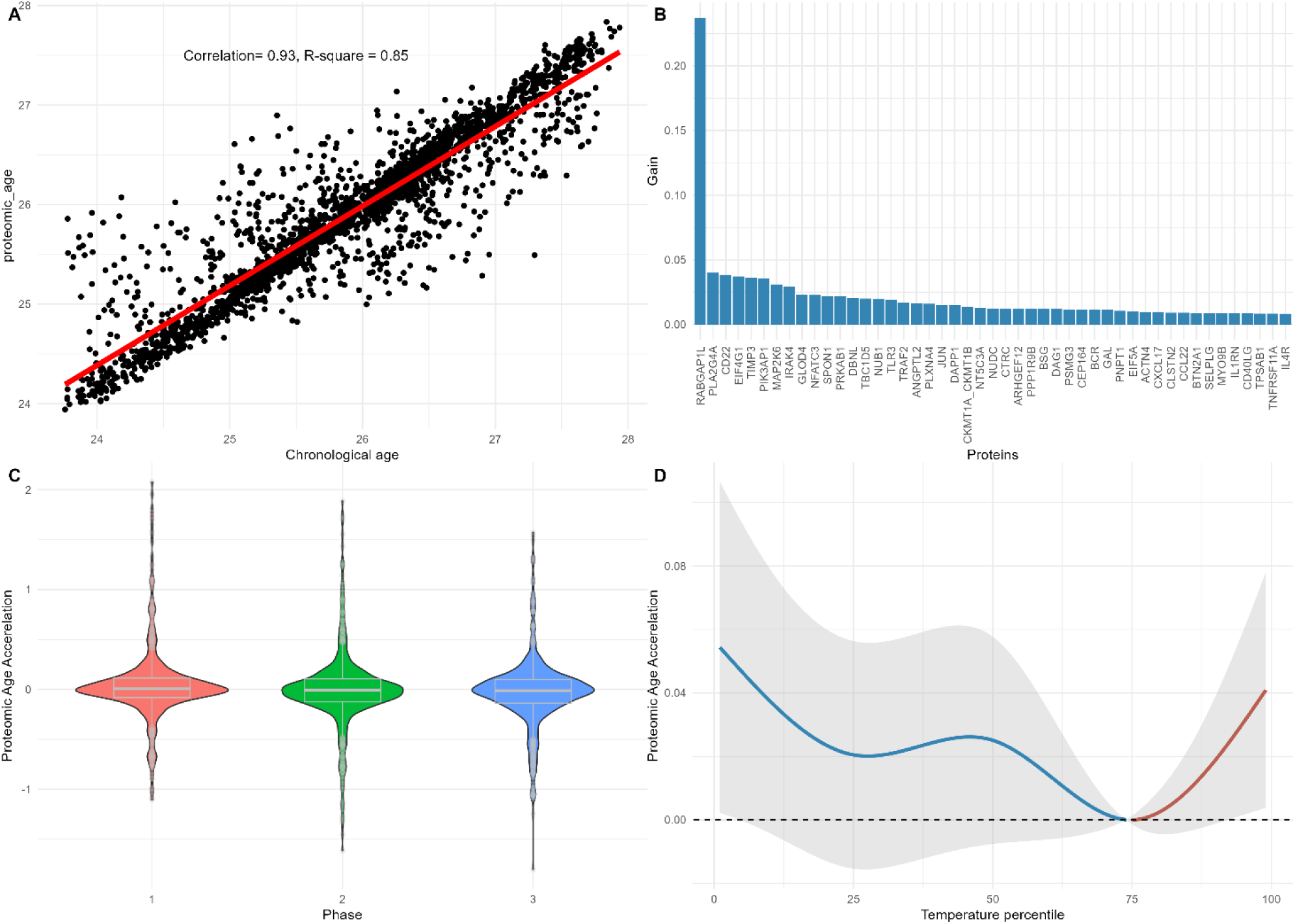
The inflammation-related proteomic aging clock and its association with temperature exposure. (A) The scatterplot between the chronological age (x-axis) and the inflammation-related proteomic age (y-axis). The solid red line represents the linear fit between the chronological age and proteomic predicted age. (B) The importance of the 47 inflammation-related proteins selected to predict the chronological age. (C) The distribution of proteomic age acceleration calculated as protein predicted age minus chronological age. (D) The exposure-response curve between temperature exposure and proteomic age acceleration derived from the linear mixed-effect model with distributed lag-nonlinear models. The model was adjusted for age, sex, long time trend (nature spline term with six degrees of freedom), phases, day of the week, season (warm: April to September; cold: October to March), history of SARS-CoV-2 infection and vaccination at the time of biosampling.

## Discussion

In this repeated measurement study design within a Swedish population-based prospective cohort of young adults, we found that short-term ambient temperature exposure was significantly associated with various inflammation-related blood proteins. These proteins were significantly associated with cardio-metabolic phenotypes in our cohort and in the UK Biobank. We also observed significant modification of the cold effect on blood proteins by sex, smoking, asthma, and air pollution levels. Furthermore, cold and heat exposure were associated with significant proteomic age acceleration.

Previous evidence on temperature exposure and inflammation-related proteins is restricted to typical inflammation markers^7,9–12^ such as interleukin-6 or C-reactive protein. To our knowledge, only one study reported cross-sectional associations between low temperature and 71 subclinical inflammation-related proteins (also measured using the PEA method) in an adult cohort^8^. Five of these proteins overlapped with the findings from our study (AXIN1, CD40, CSF1, DNER, IL18). However, our study expands upon previous research by utilizing a repeated-measurement design, which offers better control for confounding factors and provides a more robust assessment of temporal relationships. In addition to confirming previous findings, we present novel associations for several other inflammation-related proteins, suggesting a broader impact of ambient temperature exposure on human health. Among proteins significantly associated with FEV1 or FVC (**Figure 3**), higher levels of certain proteins resulting from cold exposure were linked to lower lung function. Cross-sectional analysis in Phase 2 also suggests a relationship between cold exposure and lower lung function measured by spirometry (**Supplemental Figure 7**). However, prior studies have yielded mixed results on the relationship between temperature and lung function, possibly due to the differences in population and exposure range. For instance, in the Framingham Heart Study, an increase in weekly temperature before spirometry measurement was associated with lower FEV1 during winter and spring^29^. Similarly, analyses of 19,128 middle-aged general participants^30^ and 4,992 asthmatic adults^31^ from China reported an inverse J-shaped exposure-response curve, where both low and high ambient temperatures were associated with reduced lung function parameters comparing with the referent temperature (around 90 percentile of the local temperature distribution), including FEV1, FVC, and peak expiratory flow (PEF). On the contrary, two studies conducted among elderly patients with chronic obstructive pulmonary disease (COPD)^32,33^ reported no significant associations between temperature and lung function. Notably, most previous studies focused on elderly populations or patients with asthma or COPD and our findings contribute to the growing body of evidence by demonstrating that the impact of extreme temperatures can also be observed among young adults. Furthermore, altered levels of inflammatory proteins may play a role in this association.

SCGB1A1, also known as the club cell secretory protein CC16, was associated with a higher FEV1/FVC ratio, lower blood pressure, and HbA1c, as confirmed by the UKB-PPP data. CC16 is an anti-inflammatory protein highly expressed in the lung and is a potential protective factor against impaired lung function^34^ and the progression of lung diseases^35,36^. We found short-term exposure to low temperatures associated with elevated levels of CC16 within individuals, which aligns with the evidence from panel studies^37,38^ and can partly be explained by the increased airway epithelial permeability due to short-term cold exposure. One previous study also reported an association between CC16 and blood pressure^39^, although evidence remains scarce, and further investigations are needed.

We observed significantly stronger association estimates of temperature on nine proteins (TNFRSF13B, SPRY2, SPINT2, PREB, NBN, LAP3, FXYD5, CTSC, ADGRE2) in males compared with females and three proteins vice versa (EIF5A, SKAP2, CXCL1). However, the role of sex in modifying the temperature-inflammation biomarker^8^ or temperature-mortality/morbidity^40,41^ outcomes was not consistent, which merits further investigation. The association estimates were stronger among participants with asthma for three proteins (ADGRE2, NBN, CTSC), in line with previous studies that cold air temperature exposure triggers bronchoconstriction and airway hyperresponsiveness in asthmatics patients^42^ and increased airway inflammation in an animal model^43,44^. Interestingly, we also observed interactions between air pollution and cold temperature: the estimates of cold on BCR and ANGPTL2 were significantly stronger in the presence of higher PM_2.5_ exposure, which is consistent with the previous epidemiological evidence on the synergistic effect of air pollution and temperature on health outcomes^45–47^.

While the clinical implications of these protein perturbations require further investigation, by applying a machine learning method, we built a proteomic aging clock within the study population. We found that exposure to both peak cold and heat was associated with accelerated biological aging. Accelerated biological aging has been associated with increased risk of multiple chronic diseases, multimorbidity and all-cause mortality^48^, indicating that extreme temperature exposure does not only affect the abundance of perturbating certain circulating proteins but has an overall adverse impact on human health. This is particularly important in the context of a changing climate as well as a worldwide aging population.

Our observations also have possible clinical implications: 1) Ambient extreme temperature exposure seems to impact levels of inflammation-related blood proteins, and these proteins may serve as potential susceptibility biomarkers; 2) Some of these proteins were associated with cardiometabolic and pulmonary phenotypes, indicating the possible broad impact of ambient temperature exposure on human health; 3) These proteins might be useful to help identify potentially vulnerable sub-populations; 4) Interactions between air pollution exposure and temperature was observed, highlighting the importance to adopt the exposome approach which takes the multiple environmental exposures into account for future studies.

The strengths of our study include the longitudinally repeated self-sampling design, a relatively large sample size, an extensive number of explored inflammation-related blood proteins, and the use of high-resolution spatial-temporal temperature modeling. One potential limitation is that the clinical phenotypes and seasonal diseases were only assessed in Phase 2. At the same time, the longitudinal design of the association testing between temperature and proteins precluded us from linking the temperature exposure to phenotypes using proteins as mediators. While an estimate of land-surface temperature in 1 km grids is highly relevant from a population and planning perspective, it is unclear how close it relates to true individual exposure. We identified weaker effects from high-temperature exposure on inflammation-related proteins than low-temperature. This could be explained by the fact that this study was conducted in a relatively cold climate zone. Additionally, no blood sampling was performed during the summer holidays in July, the warmest month in Sweden; thus, we could not fully capture the biological effect of peak heat, preventing us from generalizing our findings to a global setting. The data was collected during the COVID-19 pandemic, and the rate of COVID-19 vaccination was significantly higher during Phase 3 follow-up. This might be a potential confounder as temperature exposure was also lower in Phase 3 than in the other two follow-ups. However, phases were adjusted as one fixed-effect term in the main model. The temperature associations with proteins remained significant in the sensitivity analysis restricted to only Phases 1 and 2, where very few individuals were vaccinated. This indicates that it is unlikely that systematic differences between follow-ups were the main driver of the current findings. We also acknowledged the existence of possible residual confounding factors, such as upper airway respiratory infections, which are more common in the cold seasons. Future studies may benefit from integrating the proteomics with other omics layers to gain further insights^49^.

It should be noted that the blood samples were collected from the fingertips. All participants were asked to clean their hands with warm water for a few minutes, reducing the possibility that the blood collected from the fingers had different thermal conditions from the circulatory system. We further investigated the influence of normalization on the outcomes and used an alternative batch correction approach. As shown in **supplementary figure 8-9**, the predominant number of observations remained similar for different normalization methods, suggesting that our findings were robust and insensitive to data processing used in other projects^28^. We have not measured all possible temperature-sensitive blood proteins because we focused on circulating inflammation-related proteins. Although the repeated measurement design has generally higher internal validity, our findings are based on a young Swedish population and may not be generalizable to other populations.

In conclusion, our longitudinal study suggests that ambient temperature exposures are associated with significant changes in inflammation-related blood proteins. These findings highlight the potential role of temperature in influencing cardio-metabolic health and lung function in young adults. This has a likely long-term impact on disease risk^49^. Finally, our findings shortlist circulating proteins to identify subpopulations more vulnerable to cold or heat temperature exposures.

## Supporting information

Supplemental Results

## Acknowledgments

The study received funding from the Swedish Research Council (grant no. 2020-01886, 2022-06340), the Swedish Research Council for Health, Working Life and Welfare (FORTE grant no.2017-01146, no.2023-01213), the Swedish Heart-Lung Foundation, Karolinska Institute (no. 2022-01807) and Region Stockholm (ALF project for cohort and database maintenance). We acknowledge the Knut and Allice Wallenberg Foundation for supporting the Human Protein Atlas. We thank the children and parents participating in the BAMSE cohort and all staff involved in the study through the years. We thank SciLifeLab’s Affinity Proteomics Units in Stockholm and Uppsala for supporting the DBS sample processing and the Olink analysis.

## Author contributions

Z.Y., J.M.S., and E.M. were responsible for the conceptualization of the study. S.B. and A.B. were involved in the data collection. F.N. was responsible for preparing the exposure data. Z.Y., J.Z., P.L., and M.S. were responsible for the statistical analysis plan. Z.Y. performed the statistical analysis, visualized the results, and wrote the original first draft. E.M., A.B., I.K., and A.S.M. acquired the funding. All authors contributed to the interpretation of the results, revision, and edition of the manuscript.

## Data availability statement

The datasets analyzed during the current study are not publicly available due to legal and ethical regulations, but the derived data supporting the findings of this study are available from the PI of the BAMSE cohort (Professor Erik Melén) upon reasonable request.

