## Supplemental Results for "Impact of ambient temperature exposure on inflammation-related proteins: a repeated measurement study in the BAMSE cohort"

* shared last authors

**Supplemental Table 1** Comparison of the basic characteristics of the included participants and all participants at the 24-year follow-up and participants contributed to COVID-19 follow-up Phase 1

**Supplemental Table 2** Full list of the proteins significantly associated with temperature exposure

**Supplemental Table 3** Associations between inflammation-related proteins linked to temperature and clinical phenotypes in the BAMSE study and UKB-PPP

**Supplemental Table 4** Association estimates between cold and inflammation-related proteins in the main and sensitivity analysis (in Excel format)

**Supplemental Figure 1** Timeline of the COVID-19 follow-ups of the BAMSE study and Flow chart of the included blood samples

**Supplemental Figure 2** Spearman correlations between temperature, air pollution levels, and relative humidity.

**Supplemental Figure 3** Exposure-response curves for proteins significantly associated with temperature

**Supplemental Figure 4** Sensitivity analysis on the association between extreme cold and inflammation-related proteins

**Supplemental Figure 5** Overlapping inflammation-related proteins associated with lung function, blood pressure, and HbA1c

**Supplemental Figure 6** Results of the protein-protein interaction network analysis on all proteins associated with temperature

**Supplemental Figure 7** Cross-sectional association results between short-term exposure to temperature (lag0-4) and lung function, blood pressure, and HbA1c

**Supplemental Figure 8** Comparison of association estimates of extreme cold exposure with top20 inflammation-related proteins using the plate control normalization and intensity normalization method

**Supplemental Figure 9** Comparison of association estimates of extreme cold exposure with all the inflammation-related proteins using the plate control normalization and intensity normalization method

**Supplemental Table 1** Comparison of the basic characteristics of the participants included in this study,all participants of the 24-year follow-up, and subjects that contributed to the Phase 1 of the COVID-19 follow-up.

| Characteristics | Included participants (N=807) | All participants that attended the 24-year clinical examination (N=2270) | Phase 1 questionnaire (N=1664) |
| --- | --- | --- | --- |
| Sex, n (%) |  |  |  |
| Male | 291 (36.1) | 1004 (44.2) | 648 (39.4) |
| Female | 516 (63.9) | 1226 (55.8) | 996 (60.6) |
| Age at 24-year follow-up, years, mean±sd | 22.4 ± 0.4 | 22.4±0.5 | 22.4±0.5 |
| Education level at 24-year , n (%) |  |  |  |
| Elementary school/high school | 474 (58.7) | 1402 (61.8) | 997 (60.6) |
| University/college | 333 (41.3) | 859 (37.8) | 647 (39.4) |
| Occupation status, n (%) |  |  |  |
| Study | 454 (56.3) | 1201 (52.9) | 907 (55.2) |
| Empolyed | 303 (37.5) | 890 (39.2) | 628 (38.2) |
| Other | 50 (6.2) | 176 (7.8) | 109 (6.6) |
| Body mass index, kg/m2, mean±sd | 22.8±3.8 | 23.1±3.9 | 23.0±3.8 |
| Current smoking, n (%) | 142 (17.6) | 457 (20.1) | 311 (18.9) |
| Asthma, n (%) | 110 (13.6) | 333 (14.7) | 241 (14.7) |
| Lung function at 24-year follow-up |  |  |  |
| FEV1, L | 3.9±0.8 | 4.0±0.8 | 4.0±1.0 |
| FVC, L | 4.7±1.0 | 4.9±1.1 | 4.9±1.1 |

**Supplemental Table 2** Full list of the proteins significantly associated with temperature exposure.

| Gene name | Protein Name | Function^*^ | FDR adjusted P values |
| --- | --- | --- | --- |
| NFATC1 | Nuclear factor of activated T-cells, cytoplasmic 1 | Transcription factor involved in T cell activation | 4.06E-29 |
| IL18 | Interleukin-18 | Proinflammatory cytokine involved in innate and adaptive immunity | 2.51E-27 |
| EGF | Pro-epidermal growth factor | Involved in cell growth, proliferation, and differentiation | 2.89E-20 |
| SHMT1 | Serine hydroxymethyltransferase, cytosolic | Involved in one-carbon metabolism | 1.30E-18 |
| FOXO1 | Forkhead box O1 | Involved in apoptosis, stress resistance, and metabolism | 2.71E-13 |
| NT5C3A | Cytosolic 5'-nucleotidase 3A | Involved in nucleotide metabolism | 2.71E-13 |
| CD40LG | CD40 ligand | Crucial for T cell-dependent immune responses | 2.71E-13 |
| C1QA | Complement C1q subcomponent subunit A | Involved in the classical complement pathway | 1.45E-12 |
| AXIN1 | Axin-1 | Negative regulator of the Wnt signaling pathway | 5.11E-12 |
| PREB | Prolactin regulatory element-binding protein | Regulates prolactin gene expression | 1.40E-08 |
| LTBR | Tumor necrosis factor receptor superfamily member 3 | Involved in lymphoid organ development and immune responses | 3.25E-08 |
| ATP5IF1 | ATPase inhibitor, mitochondrial | Regulates mitochondrial ATP synthesis | 3.39E-08 |
| EIF5A | Eukaryotic translation initiation factor 5A-1 | Promotes translation elongation and termination | 4.34E-08 |
| METAP1D | Methionine aminopeptidase 1D | Involved in protein modification and processing | 8.74E-08 |
| CTSC | Dipeptidyl peptidase 1 | involved in protein degradation and immune cell activation | 1.83E-07 |
| PRDX5 | Peroxiredoxin-5, mitochondrial | involved in cellular antioxidant defense | 3.04E-07 |
| TNFRSF4 | Tumor necrosis factor receptor superfamily member 4 | involved in T cell activation and survival | 6.65E-07 |
| SPRY2 | Protein sprouty homolog 2 | Negatively regulates receptor tyrosine kinase signaling | 7.00E-07 |
| PSMG3 | Proteasome assembly chaperone 3 | Involved in proteasome assembly | 7.29E-07 |
| FXYD5 | FXYD domain-containing ion transport regulator 5 | Modulates Na+/K+-ATPase function | 9.46E-07 |
| FIS1 | ATP synthase subunit beta | Role in ATP production within mitochondria | 1.28E-06 |
| BSG | Basigin | Involved in cell adhesion and signaling | 1.40E-06 |
| SIT1 | Signaling threshold-regulating transmembrane adapter 1 | T-cell receptor signaling | 3.63E-06 |
| MANF | Mesencephalic astrocyte-derived neurotrophic factor | Involved in neuroprotection and ER stress response | 9.83E-06 |
| F2R | Proteinase-activated receptor 1 | Involved in platelet activation and inflammation | 1.41E-05 |
| BCR | Breakpoint cluster region protein | Involved in signal transduction and GTPase activation | 4.32E-05 |
| MAPK9 | Mitogen-activated protein kinase 9 | Involved in stress response and apoptosis | 8.07E-05 |
| TGFB1 | Transforming growth factor beta-1 proprotein | Involved in cell growth, differentiation, and immune regulation | 9.08E-05 |
| GOPC | Golgi-associated PDZ and coiled-coil motif-containing protein | Involved in intracellular protein trafficking | 0.000585764 |
| ANGPTL2 | Angiopoietin-related protein 2 | Involved in angiogenesis and inflammation | 0.000740986 |
| MATN2 | Matrilin-2 | Involved in extracellular matrix assembly | 0.000740986 |
| SPINT2 | serine peptidase inhibitor, Kunitz type 2 | Encodes a protease inhibitor protein that acts as a tumor suppressor and regulates epithelial functions. | 0.000766367 |
| TPT1 | Translationally-controlled tumor protein | Involved in calcium binding and microtubule stabilization | 0.001383389 |
| BTN3A2 | Butyrophilin subfamily 3 member A2 | Involved in T cell activation | 0.001701785 |
| HPCAL1 | Hippocalcin-like protein 1 | Primarily in calcium signaling within neurons, impacting processes related to vision and neuronal communication | 0.002257921 |
| LAMP3 | Lysosome-associated membrane glycoprotein 3 | Involved in dendritic cell function and antigen presentation | 0.002703022 |
| CHRDL1 | Chordin-like protein 1 | Involved in bone morphogenetic protein signaling regulation | 0.005040984 |
| TGFA | Protransforming growth factor alpha | Involved in cell proliferation and differentiation | 0.005761049 |
| AGRP | Agouti-related protein | Involved in appetite regulation | 0.005885621 |
| LSP1 | Lymphocyte-specific protein 1 | Involved in neutrophil transendothelial migration | 0.006679847 |
| MEPE | Matrix extracellular phosphoglycoprotein | Involved in bone mineralization | 0.008031176 |
| ADGRE2 | Adhesion G protein-coupled receptor E2 | Involved in immune cell adhesion and activation | 0.008031176 |
| TNFRSF13B | Tumor necrosis factor receptor superfamily member 13B | Involved in B cell activation and survival | 0.010228565 |
| SKAP2 | Src kinase-associated phosphoprotein 2 | Plays a vital role as an adaptor protein in immune signaling, particularly in the regulation of integrin-mediated functions in neutrophils and macrophages | 0.010228565 |
| IKBKG | NF-kappa-B essential modulator | Essential for activating NF-kappa B | 0.01154277 |
| LAP3 | Cytosol aminopeptidase | Involved in protein degradation and processing | 0.011668569 |
| IL32 | Interleukin-32 | Proinflammatory cytokine involved in innate and adaptive immune responses | 0.017452406 |
| NBN | Nibrin | Crucial for DNA double-strand break repair and maintenance of chromosome integrity | 0.019944241 |
| TRIM21 | tripartite motif-containing protein 21 | Role in linking the adaptive immune response (through antibodies) with innate immune mechanisms, primarily through its functions as an intracellular antibody receptor and E3 ubiquitin ligase | 0.022355331 |
| SIGLEC1 | Sialoadhesin | Involved in cell-cell interactions and immune response | 0.026606549 |
| CD40 | Tumor necrosis factor receptor superfamily member 5 | Pivotal receptor in the immune system, facilitating critical interactions between T cells and B cells, promoting antibody production, and regulating immune responses. | 0.026606549 |
| BANK1 | B-cell scaffold protein with ankyrin repeats | Involved in B cell receptor signaling | 0.028943906 |
| PIK3AP1 | Phosphoinositide 3-kinase adapter protein 1 | Plays a crucial role in promoting the activation of phosphoinositide 3-kinase (PI3K) signaling | 0.029586984 |
| MPIG6B | Megakaryocyte And Platelet Inhibitory Receptor G6b | Involved in the immune response, particularly in the regulation of T cell activation and function. | 0.029586984 |
| SCGB1A1 | Secretoglobin Family 1A Member 1 | Anti-inflammatory proteins that highly expressed in airway and lung | 0.336257385 |
| DAG1 | Dystroglycan 1 | Essential for the structural integrity of muscle cells and other tissues. It serves as a receptor for extracellular matrix components and is involved in cell adhesion, signaling, and the maintenance of muscle fiber integrity. | 0.040481317 |
| CXCL1 | Chemokine (C-X-C motif) ligand 1 | Acts by binding to its receptor CXCR2, promoting chemotaxis and activation of neutrophils, thereby contributing to the body's defense mechanisms against pathogens. | 0.046383676 |
| CSF1 | Macrophage colony-stimulating factor 1 | Involved in macrophage differentiation and function | 0.049179281 |

* Function according to The Human Protein Atlas (<https://www.proteinatlas.org/>).

**: P values derived from the likelihood test comparing the model with and without temperature exposure

**Supplemental Table 3** Associations between inflammation-related proteins linked to temperature and clinical phenotypes in the BAMSE study and UKB-PPP.

| Protein name | BAMSE | | | UKB-PPP | | | |
| --- | --- | --- | --- | --- | --- | --- | --- |
|  | Clinical phenotype | Association | P-value | Clinical phenotype | ICD 10 | Association | P-value |
| DNER | FEV1/FVC | Beta=0.008 | 0.003 | Pneumonia organism unspecified | J18 | OR=0.88 | 1.52E-10 |
|  |  |  |  | Unspecified acute lower respiratory infection | J22 | OR=0.92 | 2.20E-08 |
|  |  |  |  | Other chronic obstructive pulmonary disease | J44 | OR=0.87 | 1.40E-08 |
| TGFB1 | FEV1 | Beta= -0.039 | 0.026 | Other chronic obstructive pulmonary disease | J44 | OR=1.24 | 4.58E-13 |
|  |  |  |  | Pneumonia organism unspecified | J18 | OR=1.16 | 1.20E-18 |
|  |  |  |  | Respiratory failure | J96 | OR=1.27 | 1.63E-08 |
| TNFRSF13B | FEV1/FVC | Beta=-0.007 | 0.031 | Pneumonia organism unspecified | J18 | OR=1.19 | 2.51E-17 |
|  |  |  |  | Unspecified acute lower_respiratory_infection | J22 | OR=1.10 | 4.88E-11 |
|  |  |  |  | Emphysema | J43 | OR=1.31 | 8.27E-08 |
|  |  |  |  | Other chronic obstructive_pulmonary_disease | J44 | OR=1.23 | 9.67E-18 |
|  |  |  |  | Pulmonary edema | J81 | OR=1.81 | 3.91E-08 |
|  |  |  |  | Respiratory failure not_elsewhere_classified | J96 | OR=1.32 | 1.18E-11 |
| TNFRSF4 | FEV1/FVC | Beta=-0.006 | 0.024 | Pneumonia organism unspecified | J18 | OR=1.31 | 9.93E-39 |
|  |  |  |  | Unspecified acute lower_respiratory_infection | J22 | OR=1.12 | 1.81E-14 |
|  |  |  |  | Emphysema | J43 | OR=1.38 | 1.19E-10 |
|  |  |  |  | Other chronic obstructive_pulmonary_disease | J44 | OR=1.34 | 1.37E-33 |
|  |  |  |  | Bronchiectasis | J47 | OR=1.34 | 1.20E-10 |
|  |  |  |  | Pleural effusion not_elsewhere_classified | J90 | OR=1.24 | 7.49E-13 |
|  |  |  |  | Respiratory failure not_elsewhere_classified | J96 | OR=1.44 | 6.86E-19 |
| SCGB1A1 | FEV1/FVC | Beta=0.07 | 0.018 | Unspecified acute ower_respiratory_infection | J22 | OR=0.91 | 1.12E-10 |
|  |  |  |  | Emphysema | J43 | OR=0.57 | 1.10E-30 |
|  |  |  |  | Other chronic bstructive_pulmonary_disease | J44 | OR=0.58 | 7.55E-111 |
|  |  |  |  | Asthma | J45 | OR=0.82 | 8.99E-42 |
| SCGB1A1 | SBP | -1.69 | 0.001 | SBP |  | Beta=-0.07 | 3.31E-30 |
| SCGB1A1 | DBP | -0.94 | 0.030 | DBP |  | Beta=-0.06 | 1.16E-39 |
| BANK1 | DBP | Beta=0.76 | 0.048 | DBP | NA | Beta=0.05 | 3.34E-24 |
| NFATC1 | DBP | Beta=0.877 | 0.018 | DBP | NA | Beta=0.047 | 1.79E-21 |
| TNFRSF4 | DBP | Beta=-0.95 | 0.007 | DBP | NA | Beta=-0.028 | 1.15E-08 |
| C1QA | HbA1c | Beta=-0.26 | 0.018 | HbA1c | NA | Beta=-0.025 | 1.70E-07 |
| MEPE | HbA1c | Beta=-0.25 | 0.039 | HbA1c | NA | Beta=-0.395 | 1.19E-16 |
| SCGB1A1 | HbA1c | Beta=-0.295 | 0.026 | HbA1c | NA | Beta=-0.05 | 4.44E-20 |

FEV1: forced expiratory volume in 1 second, FVC: forced vital capacity, SBP: systolic blood pressure, DBP: diastolic blood pressure, HbA1c: hemoglobin A1c. UKB-PPP: UK biobank Pharma Proteomic Project

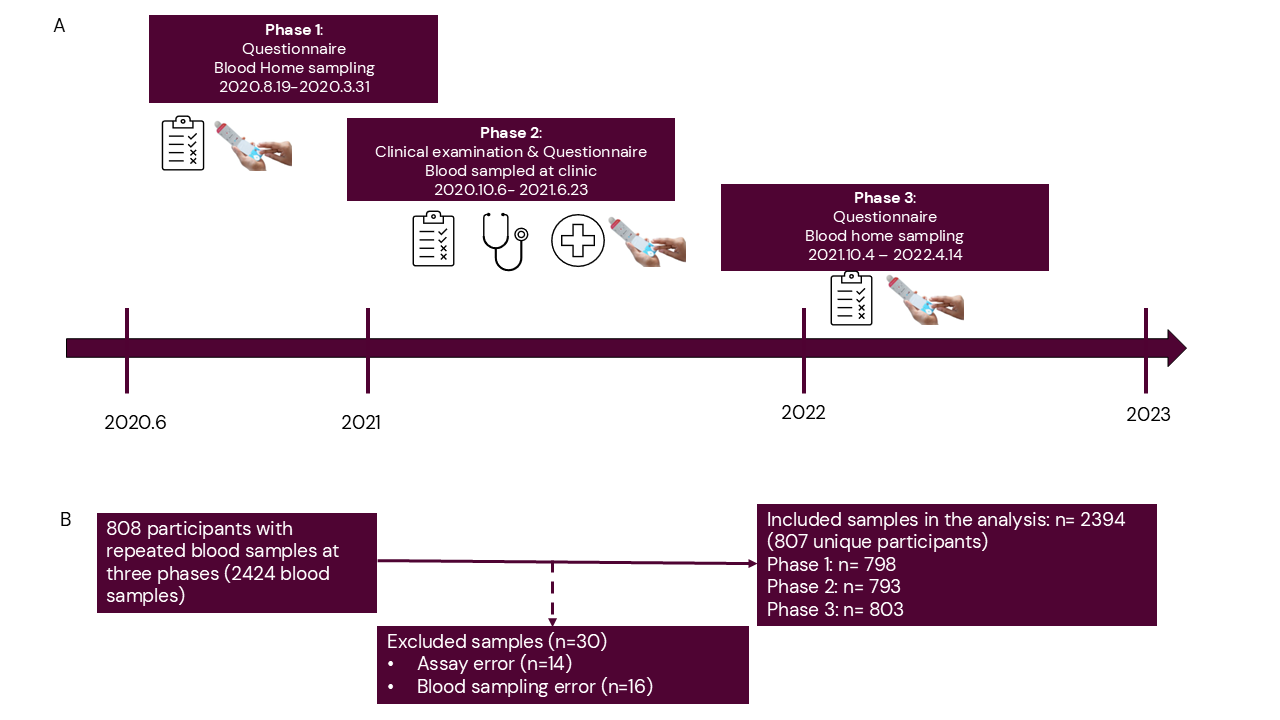

**Supplemental Figure 1**. (A) Timeline of the COVID-19 follow-ups of the BAMSE study (B) Flow chart of the included blood samples.

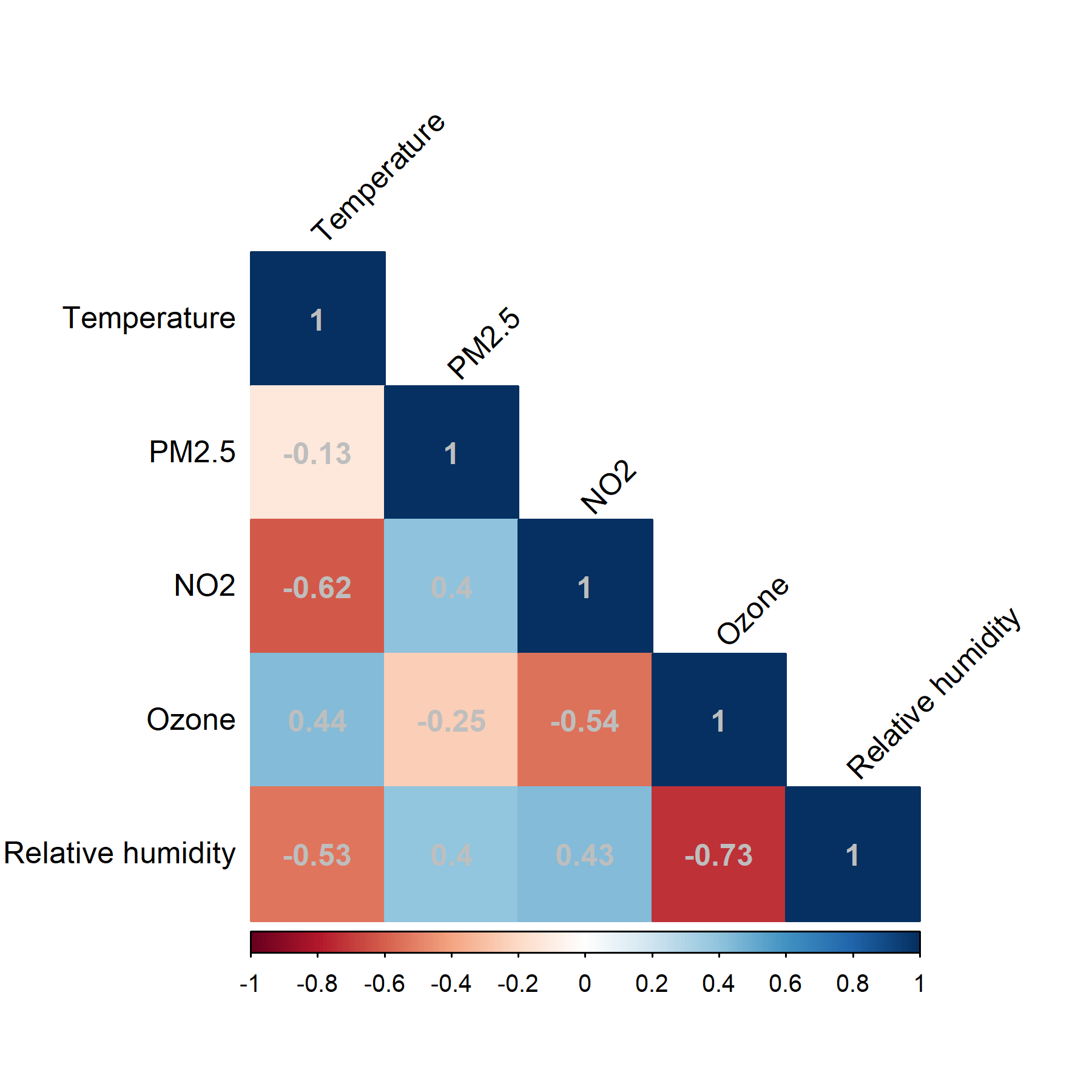

**Supplemental Figure 2.** Spearman correlations between temperature, air pollution levels, and relative humidity. PM2.5, particulate matter with diameter ≤2.5 μm; NO2, nitrogen dioxide. The correlations were calculated on the exposure of each individual.

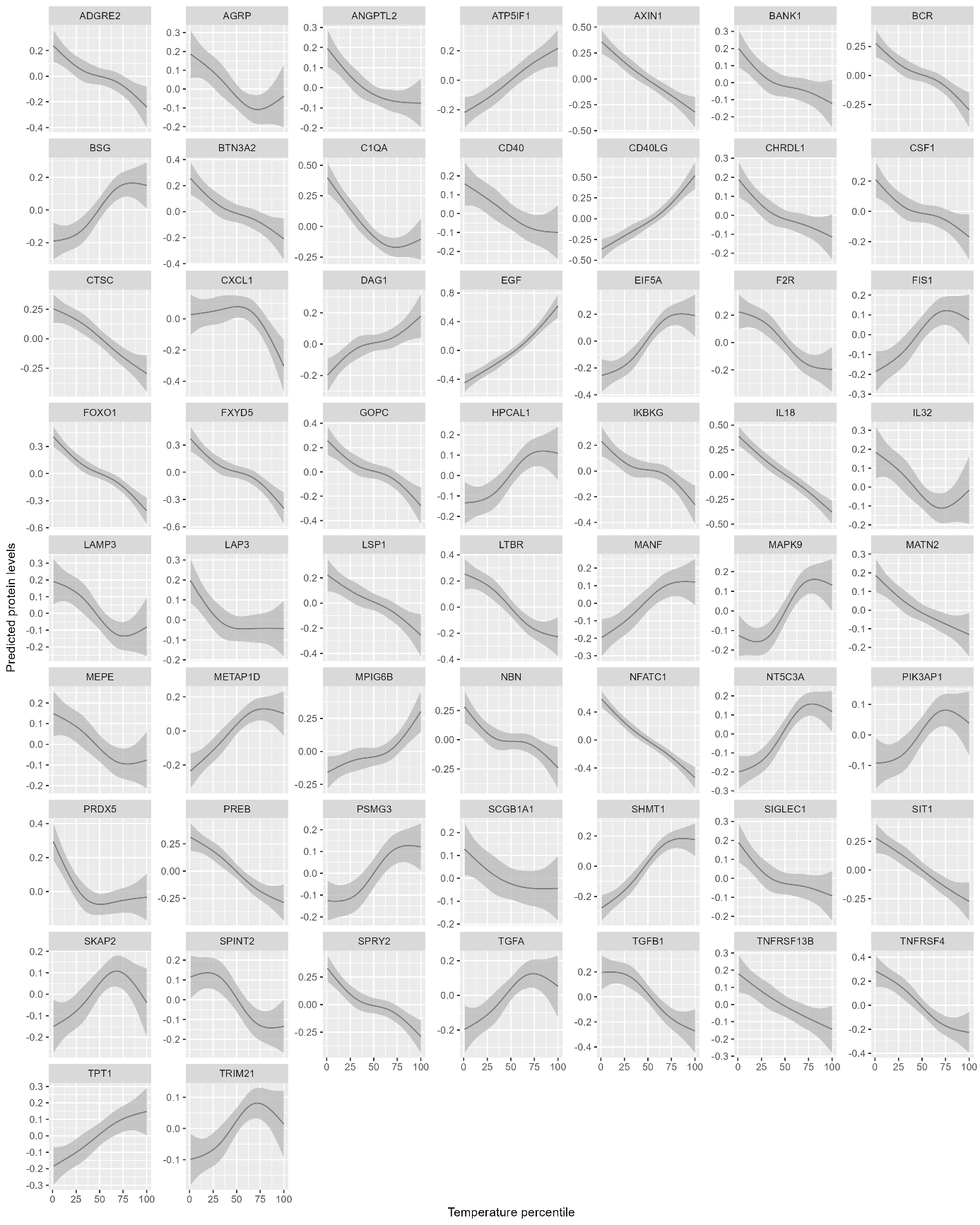

**Supplemental Figure 3.** Exposure-response curves for proteins significantly associated with temperature. The regression model was adjusted for age, sex, long-time trend (nature spline term with six degrees of freedom), phase, day of the week, season (warm: April to September; cold: October to March), history of SARS-CoV-2 infection, and vaccination at the time of biosampling. Natural splines with four degrees of freedom were applied to explore the potential non-linear relationships.

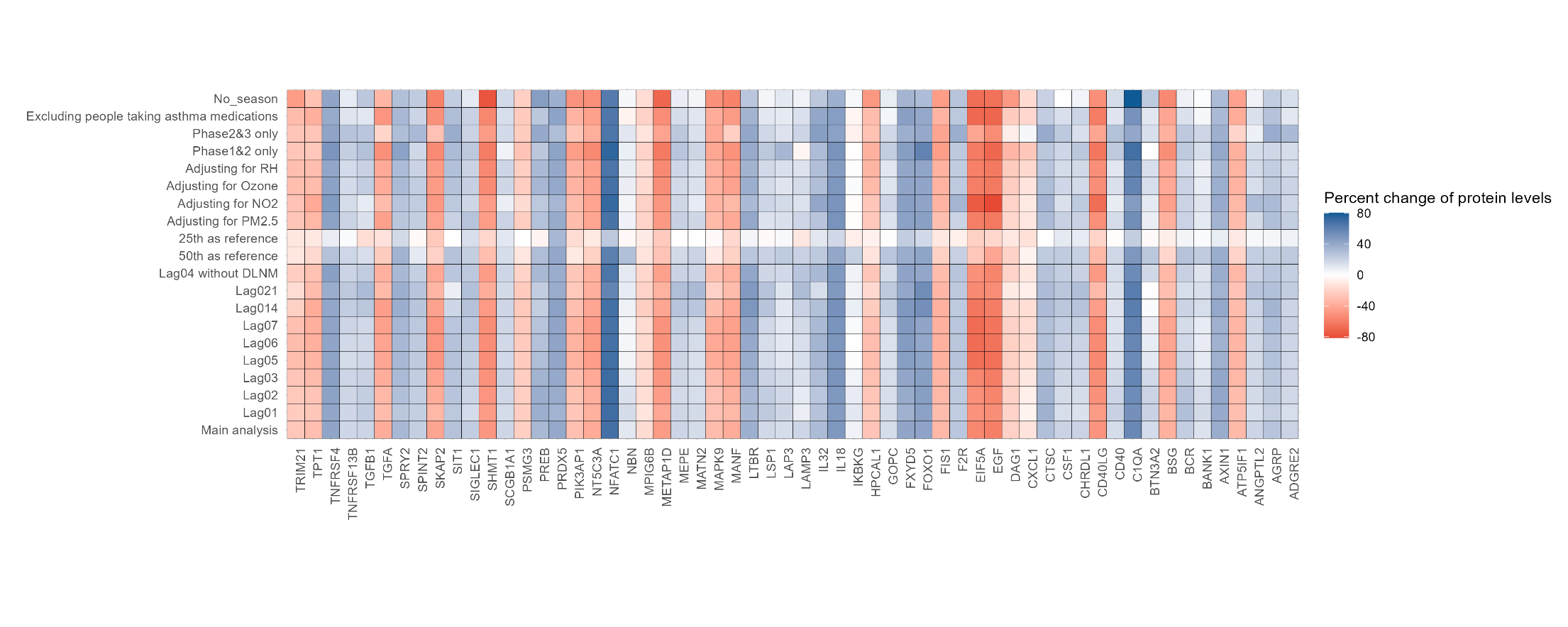

**Supplemental Figure 4** Sensitivity analysis on the association between extreme cold and inflammation-related proteins. All the results were adjusted for age, sex, long time trend (nature spline term with six degrees of freedom), phases, day of the week, season (warm: April to September; cold: October to March), history of SARS-CoV-2 infection and vaccination at the time of biosampling. Estimates presented in table format were provided in Supplemental Table 4.

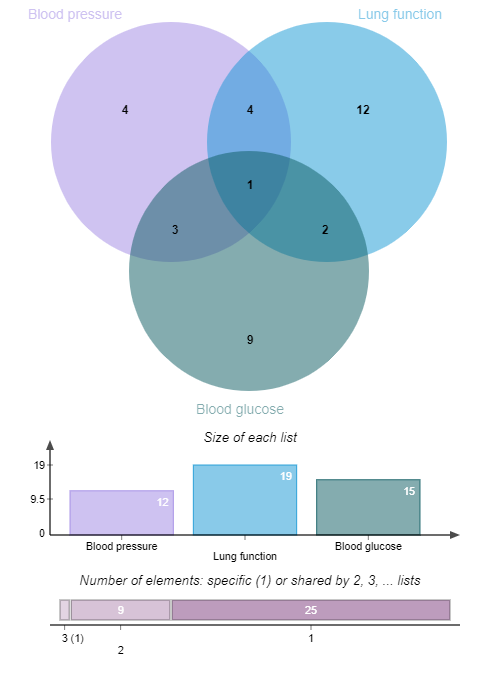

**Supplemental Figure 5.** Overlapping inflammation-related proteins associated with lung function, blood pressure, and HbA1c. Only the proteins significantly associated with temperature exposure are shown.

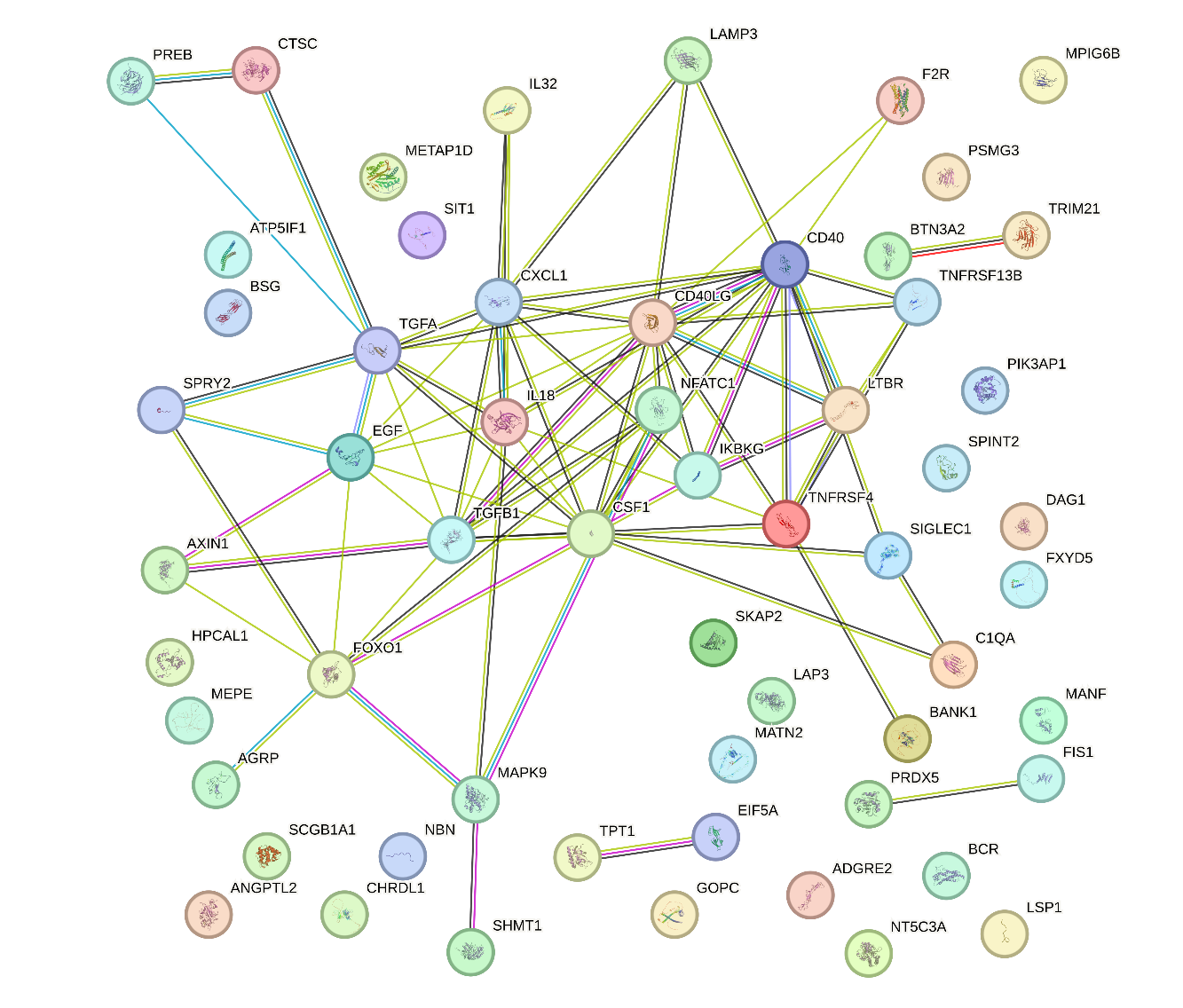

**Supplemental Figure 6** Results of the protein-protein interaction network analysis on all proteins associated with temperature

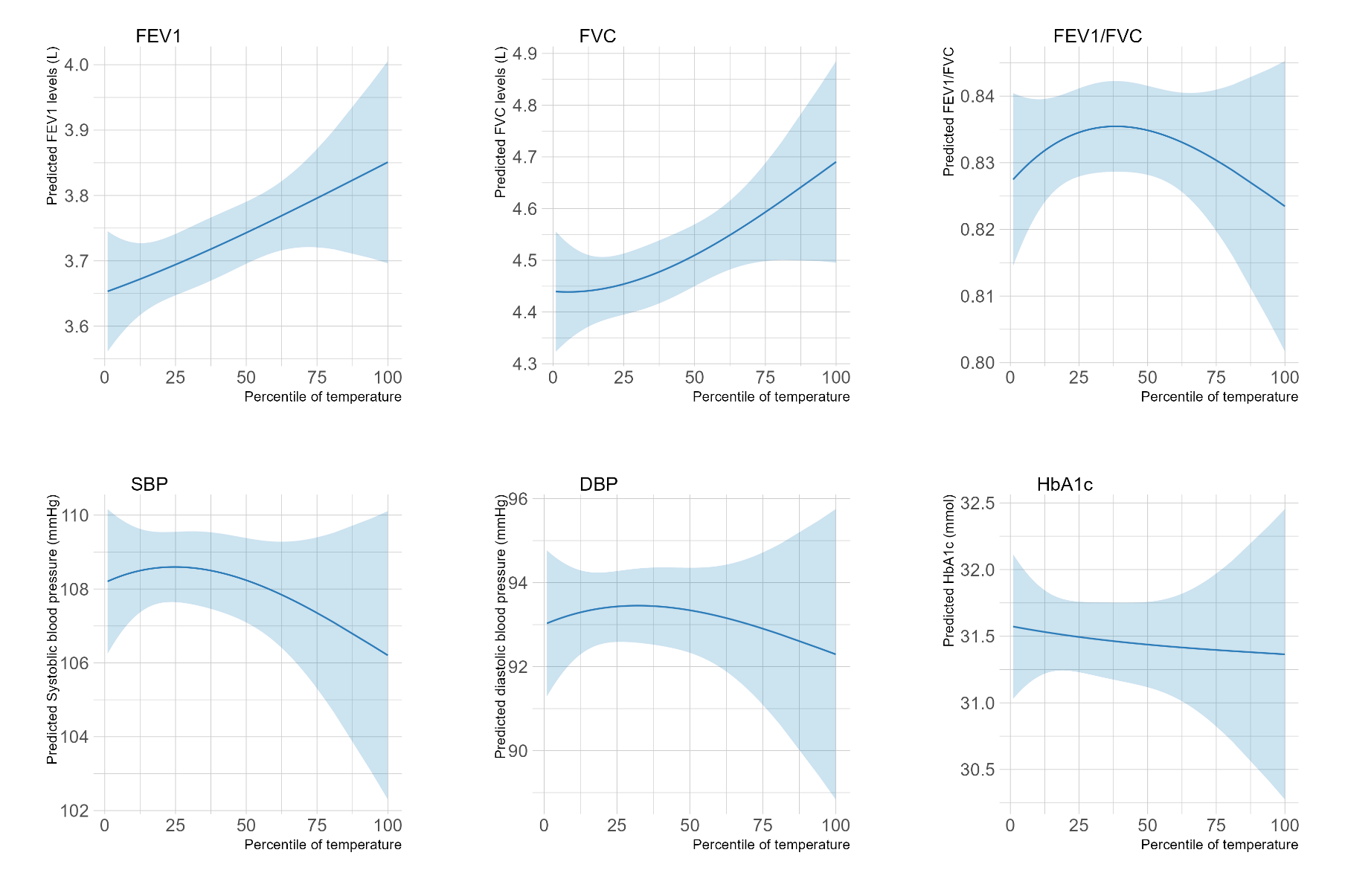

**Supplemental Figure 7.** Cross-sectional association results between short-term exposure to temperature (lag0-4) and lung function, blood pressure, and HbA1c. Nature splines with two degrees of freedom were applied to explore the potential non-linear relationship. Results were adjusted for age, sex, height (for lung function), or body mass index (for blood pressure and HbA1c). FEV1, forced expiratory volume in 1 second; FVC, forced vital capacity; SBP, systolic blood pressure; DBP, diastolic blood pressure.

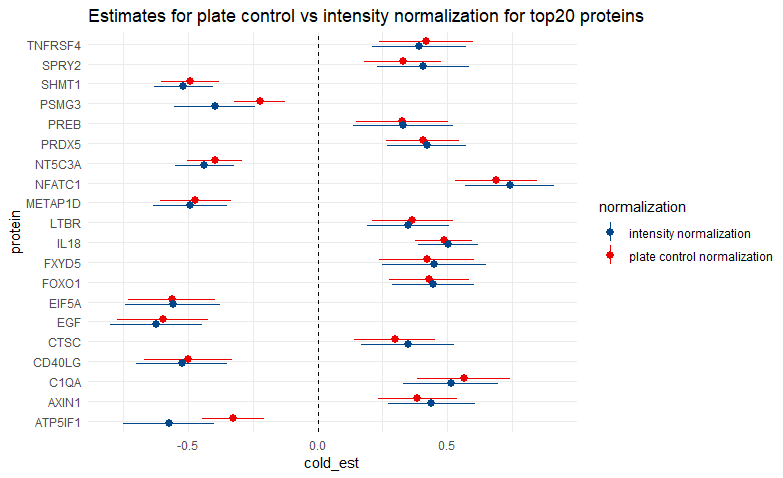

**Supplemental Figure 8** Comparison of association estimates of extreme cold exposure with top20 inflammation-related proteins using the plate control normalization and intensity normalization method. All the results were adjusted for age, sex, long time trend (nature spline term with six degrees of freedom), phases, day of the week, season (warm: April to September; cold: October to March), history of SARS-CoV-2 infection and vaccination at the time of biosampling

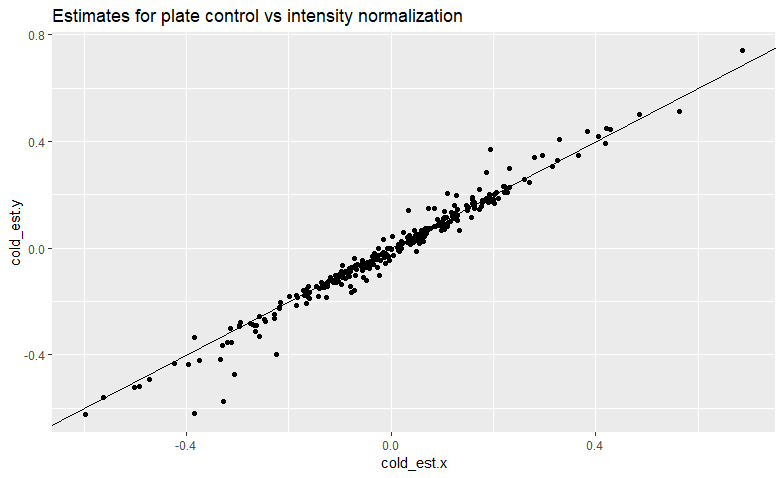

**Supplemental Figure 9** Comparison of association estimates of extreme cold exposure with all the inflammation-related proteins using the plate control normalization and intensity normalization method. All the results were adjusted for age, sex, long time trend (nature spline term with six degrees of freedom), phase of follow-up, day of the week, season (warm: April to September; cold: October to March), history of SARS-CoV-2 infection and vaccination at the time of follow-ups
